# Characterization of the Diffusion Signal of Breast Tissues using Multi-exponential Models

**DOI:** 10.1101/2020.04.27.20082271

**Authors:** Ana E. Rodríguez-Soto, Maren M. Sjaastad Andreassen, Christopher C. Conlin, Helen H. Park, Grace S. Ahn, Hauke Bartsch, Joshua Kuperman, Igor Vidić, Haydee Ojeda-Fournier, Anne M. Wallace, Michael Hahn, Tyler M. Seibert, Neil Peter Jerome, Agnes Østlie, Tone Frost Bathen, Pål Erik Goa, Rebecca Rakow-Penner, Anders M. Dale

**Affiliations:** Department of Radiology, University of California, San Diego. La Jolla, CA, USA; Department of Circulation and Medical Imaging, NTNU, Norwegian University of Science and Technology, Trondheim, Norway; School of Medicine, University of California, San Diego. La Jolla, CA, USA; Department of Physics, NTNU, Norwegian University of Science and Technology, Trondheim, Norway; Department of Radiation Oncology, University of California, San Diego. La Jolla, CA, USA; Department of Bioengineering, University of California, San Diego. La Jolla, CA, USA; Department of Radiology and Nuclear Medicine, St. Olav’s University Hospital, Trondheim, Norway

**Author notes:** Shared first authorship (Ana E. Rodríguez-Soto, Maren M. Sjaastad Andreassen). Shared last authorship (Rebecca Rakow-Penner, Anders M. Dale). **Corresponding Author:** Rebecca Rakow-Penner, MD, PhD, Department of Radiology, University of California, San Diego, 9400 Campus Point Drive #7316, La Jolla, CA 92093.

**Keywords:** Breast MRI, DW-MRI, diffusion imaging, restriction spectrum imaging, RSI

## Abstract

**Background:** Diffusion-weighted magnetic resonance imaging (DW-MRI) has demonstrated potential as an exogenous contrast-free diagnostic tool for breast cancer screening. Advanced non-Gaussian models of the DW-MRI signal provide insight into the tissue microstructure. Restriction spectrum imaging (RSI) is a mathematical framework that improves tumor conspicuity by decomposing the DW-MRI signal into separate diffusion components. The number of diffusion components and corresponding apparent diffusion coefficients (ADCs) optimal for RSI are organ-specific and determined empirically. The outputs of RSI are the signal contributions of each separable diffusion component.

**Purpose:** To understand the diffusion-weighted MRI signal of cancerous and healthy breast tissues in the context of RSI.

**Study Type:** Prospective.

**Populations:** 74 women, from two sites, with pathology-proven breast cancer.

**Field Strength/Sequence:** 3.0T

**Assessment:** The DW-MRI signal was described using a linear combination of a variable number of exponential components. The ADC for each component was estimated across all voxels from control and cancer regions of interest (ROIs), patients and sites. Once ADCs were determined, the signal contributions of each diffusion component were estimated using these fixed ADC values. Conventional ADC (mono-exponential) values were also estimated.

**Statistical Tests:** The relative fitting residual and relative Bayesian information criterion (BIC) were assessed. The signal contributions of each diffusion component were compared using analysis of variance (ANOVA) and *post-hoc* tests.

**Results:** Estimated ADCs for the bi-exponential model were D_1,2_ = 2.0 × 10^−5^ and D_2,2_ = 2.2 × 10^−3^ mm^2^/s, and D_1_,_3_=0, D_2_,_3_ = 1.4 × 10^−3^ and D_3_,_3_=10.2 × 10^−3^ mm^2^/s for tri-exponential model, which in practice is reduced to a bi-exponential model with an offset, or a three-component model. The relative fitting residuals of conventional ADC, bi-exponential and three-component models in control ROIs were 2.1%, 1.62%, and 1.03%, and 3.3%, 1.0%, and 0.3% for cancer ROIs. BIC was smaller for the three-component model, indicating an improved fitting of breast DW-MRI data compared to the bi-exponential model.

**Conclusion:** Breast DW-MRI signal was best described using a tri-exponential model. The signal contributions of the slower component in bi- or tri-exponential models were larger in tumor lesions. These data may be used as differential features between healthy and malignant breast tissues.

## Introduction

The American Cancer Society recommends that women at high risk of breast cancer receive both mammography and magnetic resonance imaging (MRI) exams yearly starting at age 30.^1^ Surveillance and diagnostic breast MRI protocols include dynamic contrast-enhanced (DCE)-MRI which require intravenous gadolinium-based contrast agents. Brain depositions of such contrast agents, with unknown sequelae, have been reported in patients with repeated exposure.^2^ In addition, some gadolinium-based contrast agents are contraindicated in patients with renal failure (glomerular filtration rate < 30ml/min/1.73m^2^) and pregnant women.^3^ Hence, there is a need for the development of contrast-free MRI protocols.

Diffusion-weighted (DW)-MRI is often a part of clinical breast imaging protocols and has demonstrated potential as a contrast-free diagnostic tool for breast cancer screening and surveillance.^4–6^ In DW-MRI, the molecular diffusion dynamics of water within tissues are probed by sensitizing the MRI signal to directionally specific molecular motion. This process yields information on the direction-dependent diffusion properties of tissues and thus allows for the characterization of tissue microstructure across different histologies.^7^ For example, the DW-MRI metric of the apparent diffusion coefficient (ADC) is reduced in breast cancer compared to benign lesions and healthy fibroglandular tissue.^8^

As more advanced models (i.e. non-Gaussian, multi-exponential) are used to describe the diffusion MRI signal, further quantitative insight of the tissue environment is provided. For instance, the diffusion-weighted MRI signal of some breast cancers displays a bi-exponential behavior, which is attributed to the presence of a fast pseudodiffusion component, related to perfusion, alongside a slow diffusion component arising from increased cellularity in tumors.^9–12^

A model that has previously demonstrated potential in improving tumor conspicuity in MRI, when evaluating disease severity and response to treatment in brain^13,14^ and prostate^15^ is restriction spectrum imaging (RSI). In RSI, an advanced linear mixture model is used to theoretically decompose the diffusion-weighted MRI signal into separate water components such as restricted, hindered and free water.^16^ For different organs, the number of discernable diffusion components and their corresponding ADCs are determined empirically. In order to apply RSI to breast cancer imaging, these parameters must be established for breast tissue. Towards this goal, the purpose of this work is to characterize the DW-MRI signals of cancerous and healthy breast tissues and understand their association with the macroscopic tissue environment of the breast.

## Methods

### MRI Data Collection

Patients from two different institutions (sites) with known breast lesions were invited to participate in this study before receiving treatment and undergo a breast MRI at 3.0T before receiving treatment. Only patients with malignant lesions confirmed by histopathologic analysis were included in this analysis. None of the patients included in this study received treatment previous to MRI examination. This study was approved by the institutional review boards from both sites. All participants provided both oral and written consent.

Data at site 1 were collected using a 3T MR750 scanner (DV25–26, GE Healthcare, Milwaukee, Wisconsin, USA) and an 8-channel breast array coil. Pulse sequence parameters: *axial DCE-MRI 3D fast spoiler gradient-recalled (SPGR) acquisition*— echo time (TE)=2.6 ms, repetition time (TR)=5.4 ms, flip angle (FA)=10°, field of view (FOV)=320 × 320 mm^2^, acquisition matrix 512 × 406, reconstruction matrix 512 × 512, voxel size 0.625 × 0.625 × 2.4 mm^3^; *axial T*_2_ *fat suppressed fast spin echo (FSE)*— TE/TR=107/4520 ms, FA=111°, FOV=320 × 320 mm^2^, acquisition matrix 512 × 320, voxel size=0.625 × 0.625 × 5mm^3^; *axial reduced-FOV echo-planar imaging (EPI) DW-MRI*— TE/TR=82/9000 ms, b-values (number of diffusion directions)=0, 500 (6), 1500 (6), and 4000 (15) s/mm^2^, FOV=160 × 320 mm^2^, acquisition matrix 48 × 96, voxel size=2.5 × 2.5 × 5 mm^3^, spectral attenuated inversion recovery (SPAIR) fat suppression, phase-encoding (PE) direction A/P, and no parallel imaging.

Data at site 2 were collected using a 3T Skyra scanner (VD13, Siemens, Erlangen, Germany) and 16-channel breast array coil. Pulse sequence parameters: *sagittal DCE-MRI 3D fast low angle shot (FLASH) acquisition*— unilateral sagittal plane; TE/TR=2.2/5.8 ms, FA=15°, FOV=180 × 180 mm^2^, acquisition matrix 256 × 256, reconstruction matrix 256 × 256, voxel size 0.7 × 0.7 × 2.5 mm^3^, generalized auto-calibrating partially parallel acquisition (GRAPPA) with acceleration factor of 2 and 36 reference lines; *sagittal T*_2_ *FSE*— unilateral sagittal plane, TE/TR=118/5500ms, FA=120°, FOV=180 × 180 mm^2^, acquisition matrix 256 × 256, voxel size=0.7 × 0.7 × 2.5 mm^3^; *sagittal EPI DW-MRI*— unilateral sagittal plane; TE=88 ms, TR=10,600 ms for 15 participants and TR=11,800 ms for 10 participants, b-values=0, 200 (6), 600 (6), 1200 (6), 1800 (6), 2400 (6), and 3000 (6) s/mm^2^, FOV=180 × 180 mm^2^, acquisition matrix 90 × 90, voxel size=2.0 × 2.0 × 2.5 mm^3^, spectral fat saturation in strong mode was used on 15 participants and SPAIR fat suppression was used on 10 participants, PE direction A/P, GRAPPA with acceleration factor of 2 and 24 reference lines.

The b=0 s/mm^2^ volumes at both sites were collected in the A/P and P/A PE directions to correct DW-MRI data for geometric and intensity distortions due to B_0_ inhomogeneities using the reverse polarity gradient (RPG) method.^17^

### Data Analysis

Analyses were performed in MATLAB R2016b (The MathWorks Inc., Natick, Massachusetts, USA). All diffusion directions for a determined b-value were averaged, and the following volumetric ROIs were manually drawn on the resulting images, informed by all available data in the exam protocol (including DCE and T_2_-weighted images): (i) control regions including either the cancer-free contralateral breast (site 1) or regions without cancer in the ipsilateral breast at least 10 mm away from the cancer lesion (site 2), (ii) whole-volume cancer lesions, and (iii) background regions. Control ROIs excluded the axillary region, large cysts (>2.5cm), and susceptibility artifacts (e.g. from surgical clips). In addition, control ROIs were initially drawn as boxes and were masked using binarized T_2_-weighted images to remove the background from the ROI. Background ROIs were drawn differently between sites because of differences in denoising and reconstruction algorithms from vendors. For site 1, background ROIs were squares of 100 voxels (10 × 10 voxels) placed in the medial region of the images. For site 2, background ROIs were drawn in the air adjacent to the breast, as the signal dropped homogeneously to zero towards the edge of the image. All ROIs were validated by two breast radiologists (R.R.P. and A.Ø.).

In addition, averaged DW-MRI data were normalized by dividing all voxels by the value of the 98^th^ percentile signal intensity value in b=0 s/mm^2^ volume to preserve T_2_ information. Data were then noise corrected as described by Gudbjartsson et al.^18^ The diffusion signal was modeled as the linear combination of multiple exponential decays:

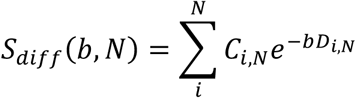

where N is the total number of exponential decays (here 2 and 3), C_i,N_ are the signal contributions of each exponential component, b are the b-values in s/mm^2^, and D_i,N_ are the ADCs of each exponential component in mm^2^/s, and D_1,N_ < D_2,N_ <D_3,N_. Global ADCs were estimated across all voxels from control and cancer ROIs, including all patients from both sites, for bi- and tri-exponential models. Model fitting was performed using a simplex search method with non-negativity constraints.^21^ To account for the different sites and imaging protocols, the fitting procedure minimized the global negative log likelihood of ADCs across all data. Initial conditions were determined using this same fitting procedure while using the signal average of both cancer and background ROIs to achieve high signal-to-noise ratio (SNR).

The relative fitting residual and relative Bayesian information criterion (BIC) were calculated for both bi- and tri-exponential models. The relative fitting residual was calculated as the difference between observed and predicted diffusion-weighted signal divided by the observed signal in all cancer and control voxels, respectively. In addition, the relative fitting residual was also calculated for conventional ADC to compare the fitting of the multi-exponential models with fixed ADCs to standard quantitative DW-MRI methods. The BIC was used because it penalizes the model’s likelihood for increasing the number of estimated parameters.^22^ Lower BIC values denote improved model fitting. However, absolute BIC values are arbitrary, therefore, we report the difference in BIC (ΔBIC = BIC_bi_ – BIC_tri_) for cancer and control voxels. After ADCs were determined, the signal contributions of each exponential component C_i,N_ were estimated for all voxels using these fixed ADC values. Two-way repeated-measures analyses of variance (RM-ANOVA) with Sidak *post-hoc* tests were used to identify differences in C_i,N_ signal contributions estimated from bi- and tri-exponential models, across diffusion components and tissues. The threshold for significance (*α*) was set at 0.05 for all analyses. Statistical analyses were performed using Prism software (version 6 for Mac OS X, GraphPad Software, La Jolla, CA, USA). All data are reported as mean ± standard deviation values.

## Results

Patients were from two sites, 57 women from site 1 and 25 women from site 2 were enrolled in this study. From site 1, eight participants were excluded from the study; six women had contralateral cancer or mastectomy and in two cases DW-MRI data were of low quality. Demographic information can be found in Table 1. Representative images of DCE, T_2_-weighted and DW-MRI (b=0 s/mm^2^) images and control, background, and cancer ROIs for both sites are shown in **Figures 1 and 2**. After noise-correction, all diffusion directions for each specific b-value were averaged (**Figure 3**). The diffusion signals of control and tumor ROIs were then fitted by bi- and tri-exponential models to estimate global ADCs.

**Table 1.**
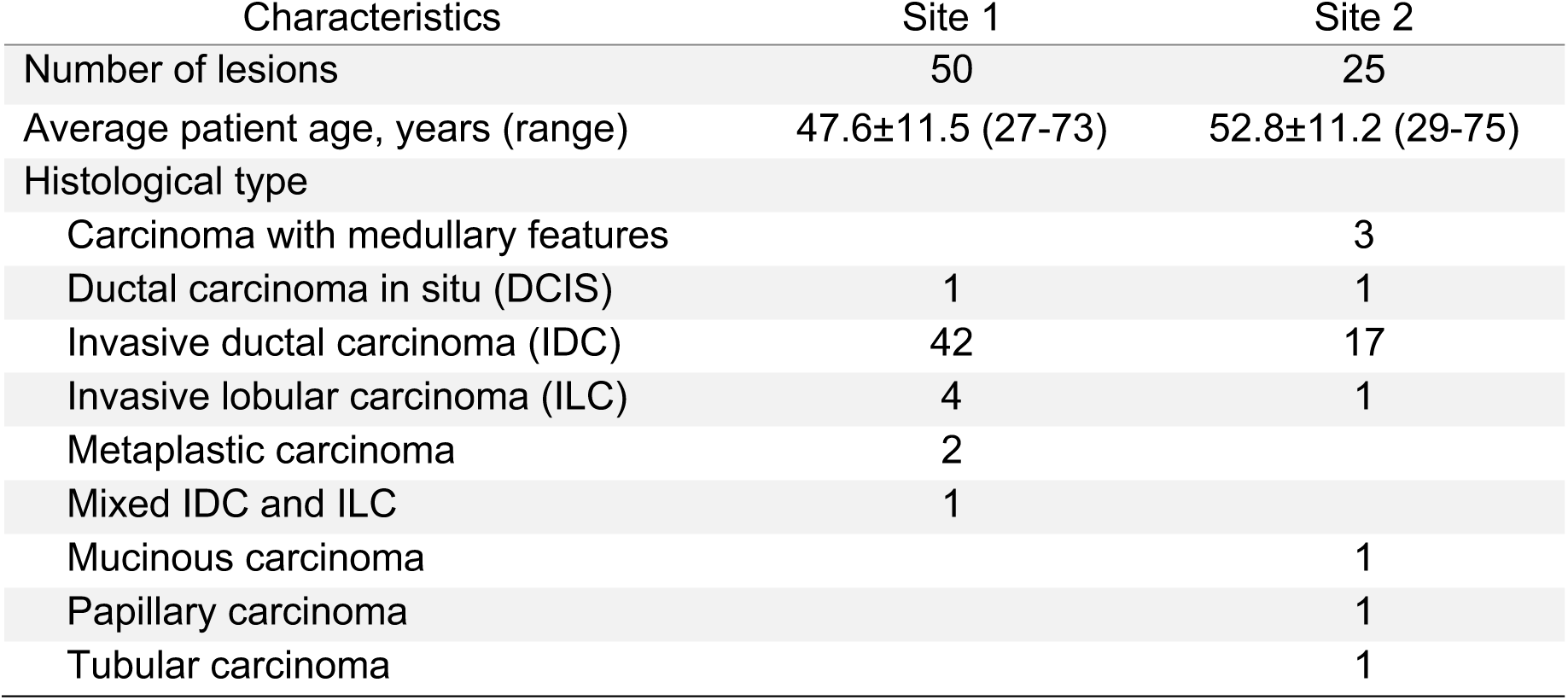
Demographic information of participants.

| Characteristics | Site 1 | Site 2 |
| --- | --- | --- |
| Number of lesions | 50 | 25 |
| Average patient age, years (range) | 47.6±11.5 (27-73) | 52.8±11.2 (29-75) |
| Histological type |  |  |
| Carcinoma with medullary features |  | 3 |
| Ductal carcinoma in situ (DCIS) | 1 | 1 |
| Invasive ductal carcinoma (IDC) | 42 | 17 |
| Invasive lobular carcinoma (ILC) | 4 | 1 |
| Metaplastic carcinoma | 2 |  |
| Mixed IDC and ILC | 1 |  |
| Mucinous carcinoma |  | 1 |
| Papillary carcinoma |  | 1 |
| Tubular carcinoma |  | 1 |

**Figure 1.**
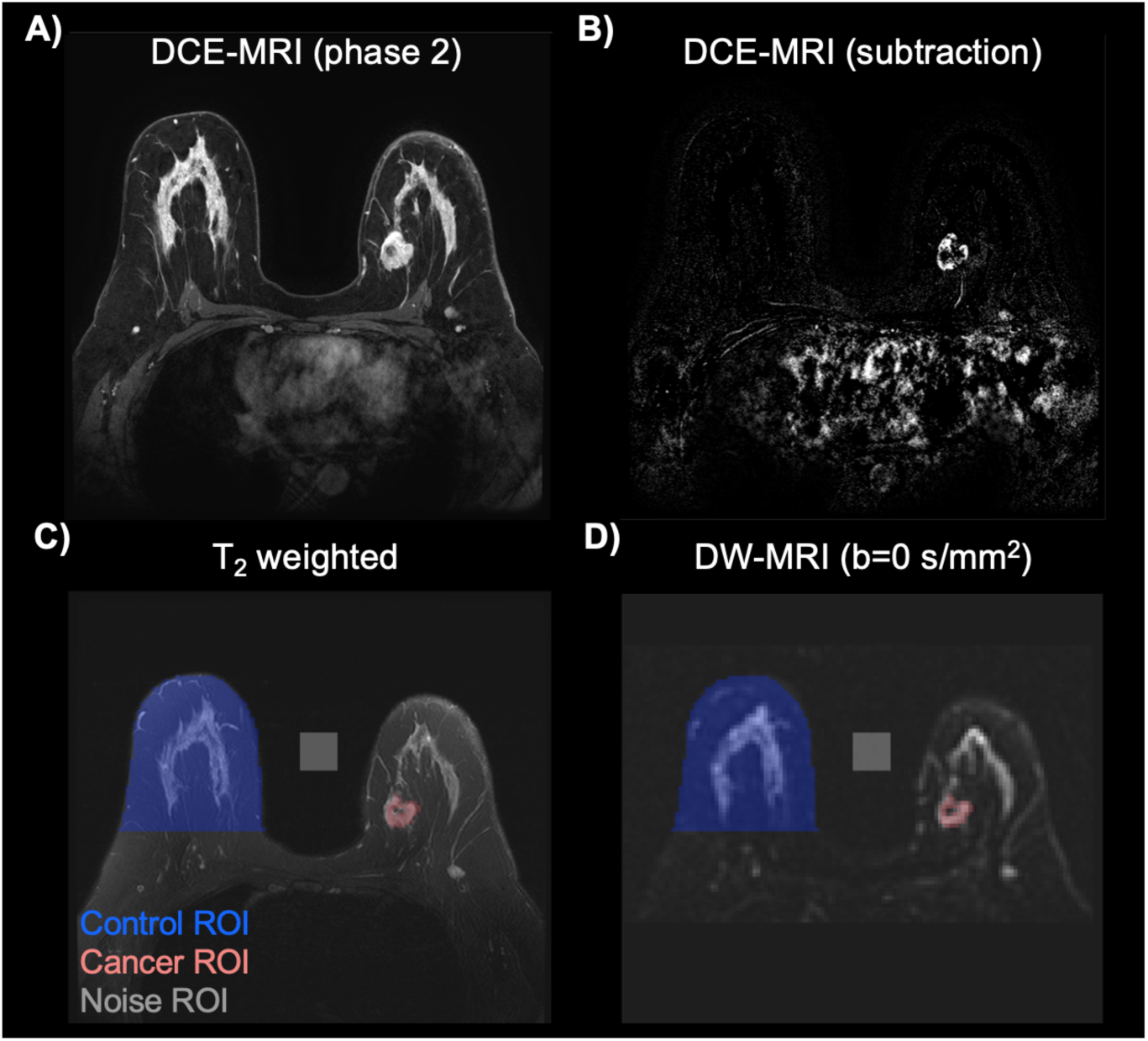
Example images of A) dynamic contrast-enhanced (DCE) MRI, B) DCE pre/post contrast subtraction, C) T_2_-weighted, and D) DW-MRI b=0 s/mm^2^ volumes from site 1. Overlaid regions of interest (ROIs) are control (blue), cancer (red), and noise (grey), respectively.

**Figure 2.**
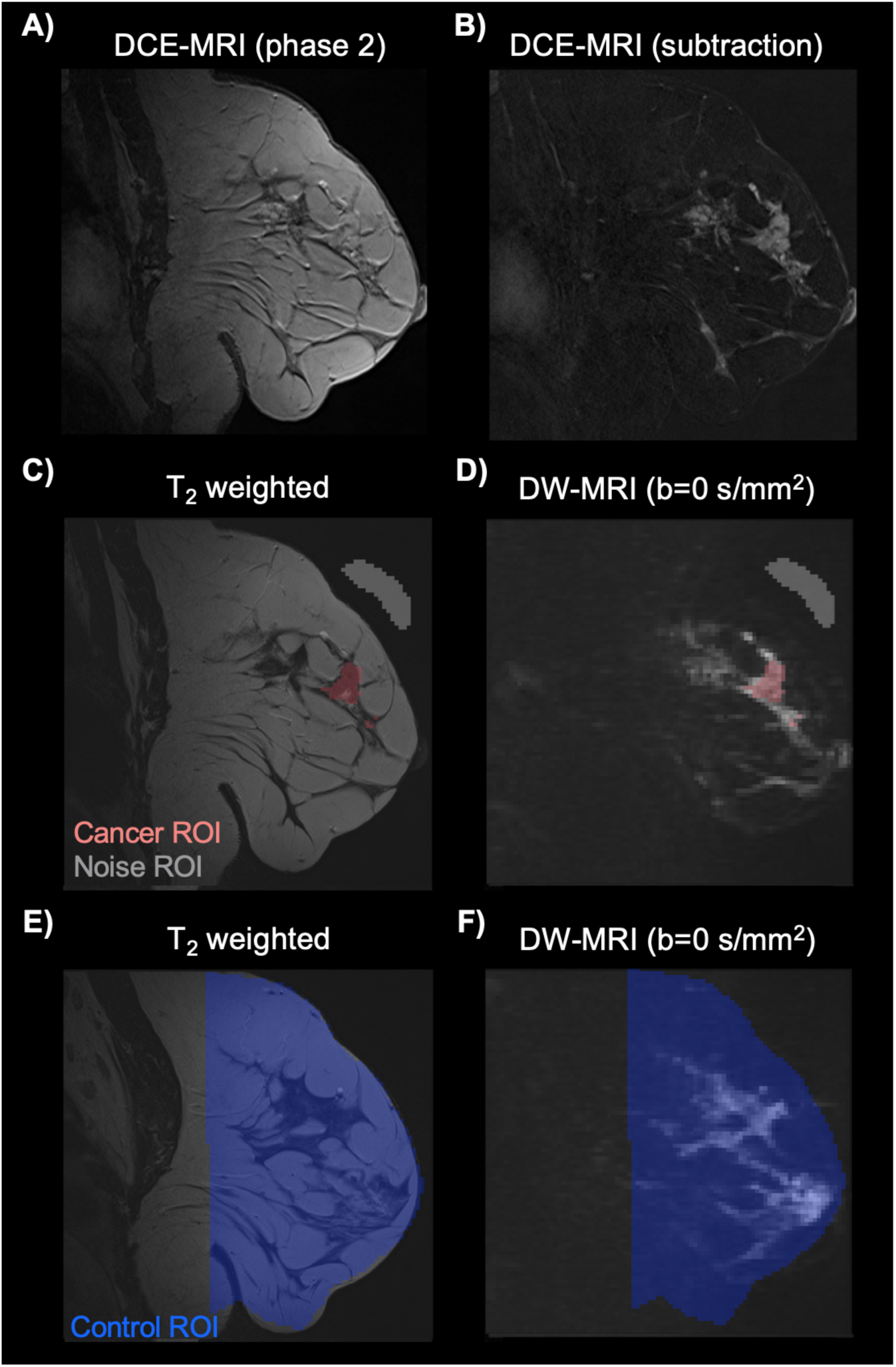
Example images of A) dynamic contrast-enhanced (DCE) MRI, B) DCE pre/post contrast subtraction, C) and E) T_2_-weighted, and F) and D) DW-MRI b=0 s/mm^2^ volumes from site 2. Overlaid regions of interest (ROIs) are control (blue), cancer (red), and noise (grey), respectively. Note that control ROIs were placed on a different image to avoid inclusion of tumor tissue/peritumor infiltration in the control ROI.

**Figure 3.**
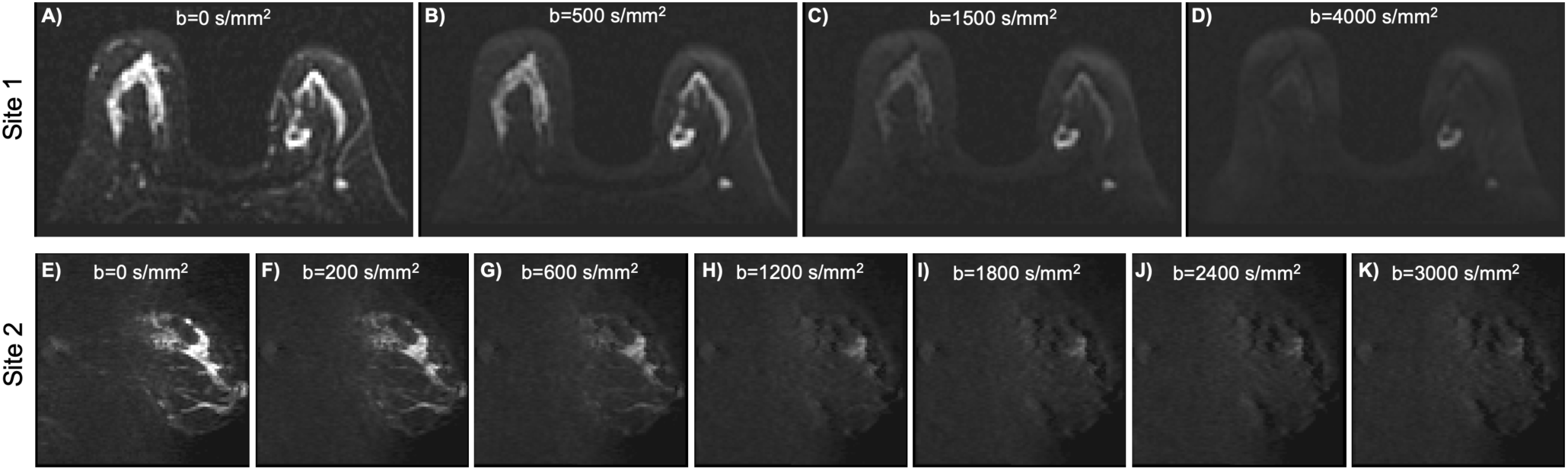
Images from diffusion-weighted magnetic resonance imaging (DW-MRI) were averaged over all diffusion directions for each b-value, and noise corrected. Representative images at different b-values and for both sites *(top and bottom rows)* are shown.

The relative fitting residuals of conventional ADC, and bi-, and tri-exponential models in control ROIs were 2.1%, 1.62%, and 1.03% of the overall signal value, while the residuals for the cancer ROIs were 3.3%, 1.0%, and 0.3%, respectively. Residuals were considerably smaller for the tri-exponential model. Similarly, a ΔBIC of 74 was estimated between the bi- and triexponential models, indicating that a tri-exponential model further improves the fitting of breast DW-MRI data.

Estimated ADCs for bi-exponential model were D_1,2_ = 2.0 × 10^−5^ and D_2,2_=2.2 × 10^−3^ mm^2^/s, and D_1,3_=1.4 × 10^−21^, D_2,3_=1.4 × 10^−3^ and D_3,3_=10.2 × 10^−3^ mm^2^/s for tri-exponential model (**Table 2**). In addition, ADCs were calculated for each site independently (**Table 2**). For the tri-exponential model, the slowest ADC is far smaller than can be quantified accurately. Thus, in this model D_1,3_ was set to 0 mm^2^/s, replacing the slowest diffusion component with a constant offset term (C_1,3_). The signal contributions C_i,N_ for control and cancer ROIs were not significantly different (p > 0.05) when estimated using D_1,3_=1.4 × 10^−21^ mm^2^/s and D_1,3_=0 mm^2^/s for the tri-exponential model. Average signal contributions of the control and cancer ROIs are shown in **Table 3**. The diffusion signal of all tissues was then described with the following three-component model:

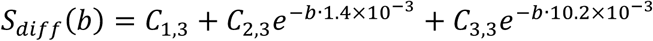

**Table 2.** Estimated apparent diffusion coefficients (ADCs) for each site independently and together in units of mm^2^/s.

| Model | Parameter | Sites 1 and 2<br>($\text{mm}^2/\text{s}$) | Site 1<br>($\text{mm}^2/\text{s}$) | Site 2<br>($\text{mm}^2/\text{s}$) |
| --- | --- | --- | --- | --- |
| Bi-exponential | $D_{1,2}$ | $2.0 \times 10^{-5}$ | $3.0 \times 10^{-5}$ | $7.1 \times 10^{-18}$ |
| | $D_{2,2}$ | $2.2 \times 10^{-3}$ | $2.1 \times 10^{-3}$ | $2.7 \times 10^{-3}$ |
| Tri-exponential | $D_{1,3}$ | $1.4 \times 10^{-21}$ | $1.4 \times 10^{-18}$ | $3.2 \times 10^{-28}$ |
| | $D_{2,3}$ | $1.4 \times 10^{-3}$ | $1.2 \times 10^{-3}$ | $1.8 \times 10^{-3}$ |
| | $D_{3,3}$ | $10.2 \times 10^{-3}$ | $9.2 \times 10^{-3}$ | $16.5 \times 10^{-3}$ |

**Table 3.** Average signal contribution of the control and cancer regions of interest (ROIs) for biexponential and three-component models, in arbitrary units (a.u.).

| Model | Parameter | Control | Cancer |
| --- | --- | --- | --- |
| Bi-exponential | $C_{1,2}$ | $0.1 \pm 0.1$ | $0.5 \pm 0.3$ |
| | $C_{2,2}$ | $0.1 \pm 0.1$ | $1.6 \pm 0.9$ |
| Three-component | $C_{1,3}$ | $0.1 \pm 0.1$ | $0.4 \pm 0.2$ |
| | $C_{2,3}$ | $0.1 \pm 0.1$ | $1.6 \pm 0.8$ |
| | $C_{3,3}$ | $0.1 \pm 0.0$ | $0.2 \pm 0.2$ |

The term “tri-exponential” is therefore replaced with “three-component” for the remainder of this paper. Control and cancer average signal contributions (C_i,N_) for the bi-exponential and three-component models are shown in **Figure 4**. In both models, statistically significant differences (p<0.0001) were found between the signal components of cancer and control ROIs. Signal contributions estimated from the bi-exponential model (C_1,2_ and C_2,2_) were different (p<0.0001) between cancer and control ROIs. Similarly, the signal contribution of the two slowest components derived from the three-component model (C_1,3_ and C_2,3_) were also different between tumor and control ROIs (p<0.0001). In addition, differences between cancer C_1,3_ and C_2,3_ were found in both models. The C_2,2_, and C_2,3_ signal contributions were significantly larger in magnitude than C_1,2_, and C_1,3_ and C_3,3_, respectively (**Figure 4**). In order to understand the inter-dependency of the multiple signal contributions in the three-component model, the average values for each subject were plotted (**Figure 5**). This showed that tumor ROIs present larger signal in both C_1,3_ and C_2,3_ than control ROIs (Figure 4A&B), while no differences between ROIs are found in C_3,3_.

**Figure 4.**
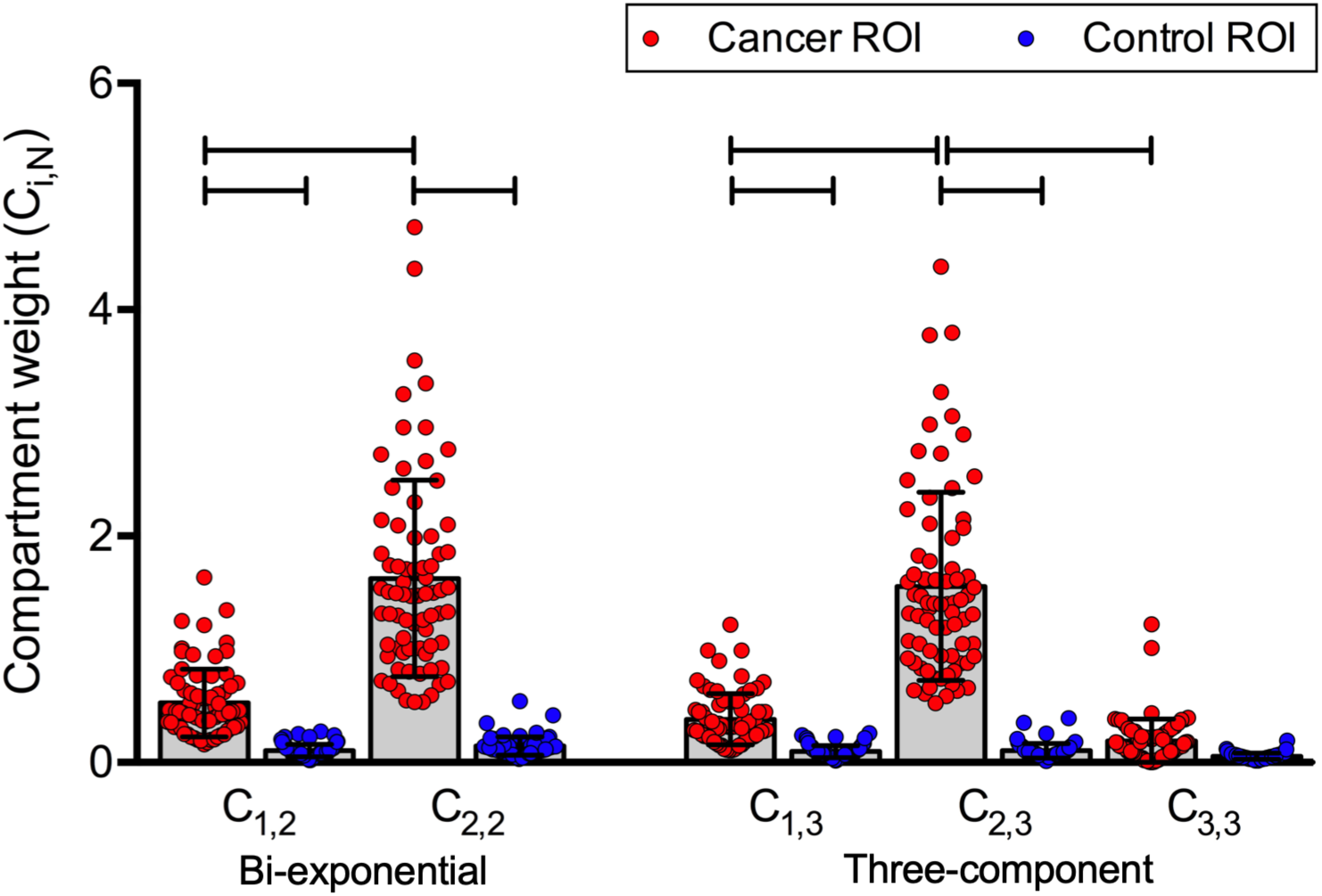
Plot of the average signal contributions of the components of the bi-exponential and three-component models within cancer (red) and control (blue) regions of interest (ROIs). In both models, the magnitude of the components of cancer and control ROIs were statistically different (p<0.0001, horizontal bars), except for the fastest component in the three-component model which is attributed to vascular flow.

Whole-volume conventional ADC and C_i,N_ maps were also generated. Representative C_i,N_ maps for each site are shown in **Figures 6 and 7**. The signal contributions from tumors and fatty tissues in both C_1,2_ and C_1,3_ (**Figures 6B,D and 7B,D**) were larger than in fibroglandular tissues. However, the signal of both tumors and fibroglandular tissues in C_2,2_ and C_2,3_ (**Figures 6C,E and 7C,E**) were larger than those of fatty tissues. The signal contribution C_3,3_ displayed information pertaining to vascular flow (D_3,3_=10.2 × 10^−3^ mm^2^/s, **Figures 6F and 7F**). In the bi-exponential model flow information appears to be contained in the fast signal contributions C_2,2_ (**Figures 6C and 7C**).

**Figure 5.**
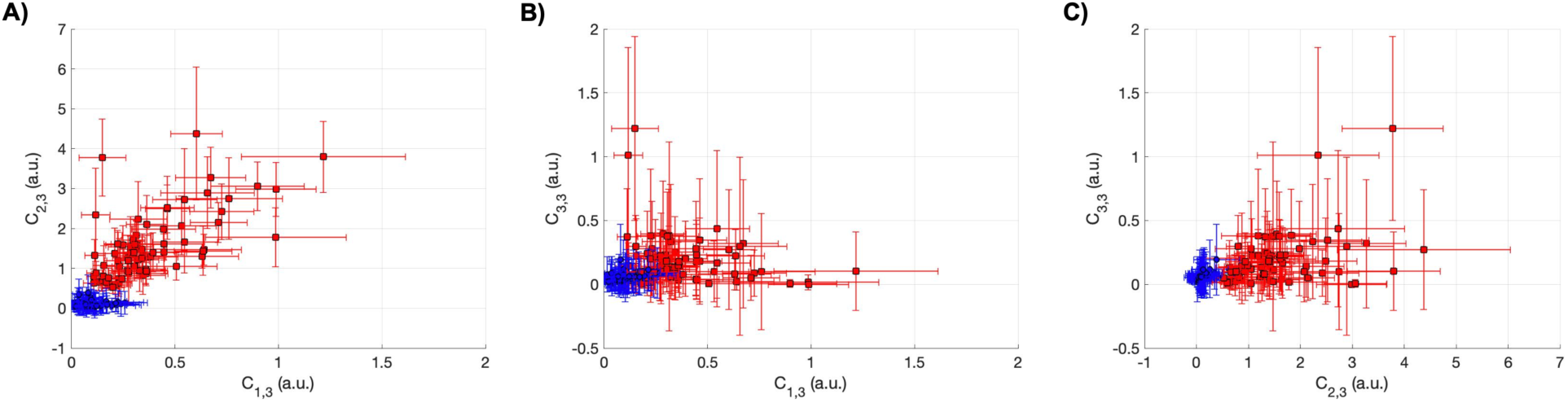
Apparent diffusion coefficients (D_i,N_) of a three-component model 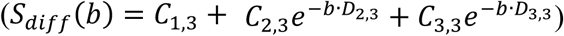 were determined by simultaneously fitting both control and cancer ROIs of both sites. Values of D_i,N_ were then fixed (D_2,3_=1.4 × 10^−3^ mm^2^/s, and D_3,3_=10.2 × 10^−3^ mm^2^/s) and used to estimate the signal contribution of each component C_i,N_. Two-dimensional plots of the magnitude of A) C_1,3_ vs C_2,3_, B) C_1,3_ vs C_3,3,_ and C) C_2,3_ and C_3,3_ are shown for control (blue circles) and cancer (red squares) ROIs. Circles and bar represent the C_i,N_ ROI average and standard deviation for each subject.

**Figure 6.**
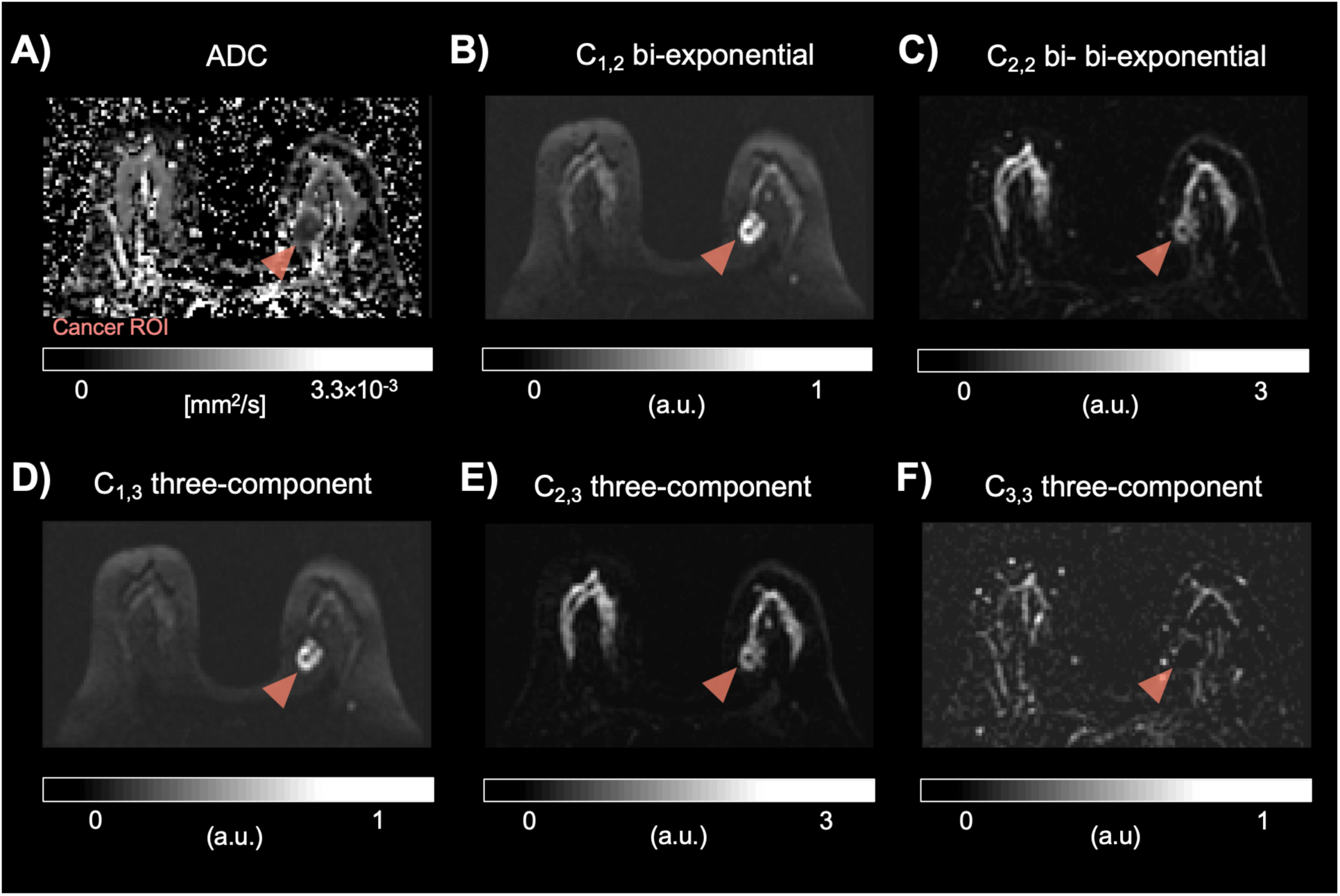
Representative images from site 1 including A) conventional ADC map, and the signal contributions (C_i,N_) of bi-exponential (B-C) and three-component (D-F) models. Arrowheads indicate tumor location. Signal contribution in tumors was higher than surrounding tissues in both C_1,N_ and C_2,N_ in both models. The compartment C_3,3_ displays vascular flow information.

**Figure 7.**
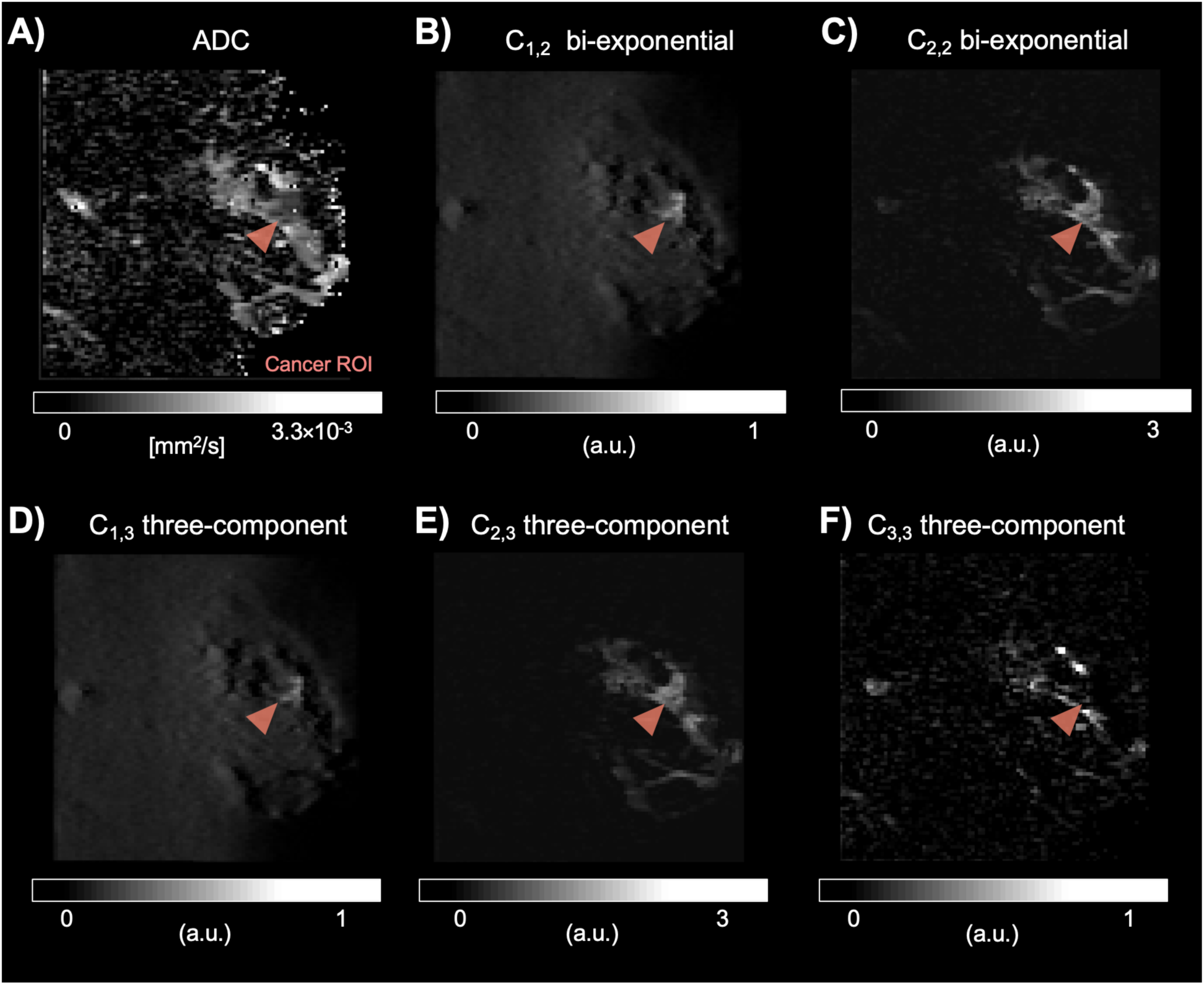
Representative images from site 2 including A) conventional ADC map, and the signal contributions (C_i,N_) of bi-exponential (B-C) and three-component (D-F) models. Arrowheads indicate tumor location. Tumor signal was higher in both C_1,N_ and C_2,N_ in both models. The compartment C_3,3_ displays vascular flow information.

## Discussion

Women at high-risk of developing breast cancer undergo annual breast MRI protocols that include DCE-MRI starting at age 30.^1^ Brain depositions of contrast agents with unknown effects have been reported in patients with repeated exposure.^2^ Further, the use of contrast agents is contraindicated in some populations, including pregnant women.^3^ Hence, there is a need for the development of contrast-free MRI protocols. Towards this goal, we describe the DW-MRI signal of healthy and cancerous breast tissues using multi-exponential models to understand the association with the macroscopic tissue environment of the breast.

Despite the quantitative nature of DW-MRI, diffusion-weighted images at high b-values are sometimes used to qualitatively inform clinical MRI exam interpretation.^4,19^ Lesions are conspicuous on these images due to the combined T_2_- and diffusion-weighted nature of the DW-MRI sequences. The combined effect of 1) lengthened T_2_,^20^ and 2) the shift in the relative size between slow and fast water diffusion components in neoplasms compared to fibroglandular tissue^19^ result in persistent signal intensity at high b-values. This effect is enhanced by the fact that breast DW-MRI data are typically fat suppressed,^21^ thereby further increasing tumor contrast-to-background. Breast RSI exploits these data collection strategies and tissue characteristics in an effort to further increase tumor conspicuity.

In the RSI framework, the diffusion-weighted MR signal at high b-values (up to 3,000–4,000 s/mm^2^) is fit to an organ-specific multi-exponential model containing fixed ADCs. Here, two and three diffusion components are used to describe the diffusion signal of breast and to determine the corresponding ADCs of each component. The main RSI outputs are the signal contributions of each component. The ΔBIC (BIC_bi_ − BIC_tri_) results suggest that a three-component model improves the characterization of the diffusion signal over a bi-exponential model. Further, the relative fitting residuals when using a three-component model are two and ten times smaller in control and cancer ROIs, respectively, compared to conventional ADC. The use of a three-component model may invite the question of data overfitting, however, the magnitudes of C_i,N_ for all tissues were dominated by two components, perhaps due to the difference in orders of magnitude of the estimated ADCs. That is, no tissue was hyperintense in all three C_i,3_ maps of the tri-exponential model.

In the three-component model, the signal contribution C_3,3_ of the fast diffusion component (D_3,3_=10.2 × 10^−3^ mm^2^/s) clearly reflects flow within vessels (**Figures 6H and 7H**). The signal contributions C_2,3_ and C_1,3_ of the two slowest diffusion components (D_2,3_=1.4 × 10^−3^ and D_1,3_=0 mm^2^/s) are then attributed to hindered and restricted diffusion, respectively. We hypothesize that the value of D_1,3_=0 mm^2^/s indicates tissue structures that exhibit restricted diffusion (reflected as persistent signal intensity at high b-values) and have minimal diffusion at the diffusion times used here.^22^ This magnitude of both flow and hindered diffusion components (C_2,3_ and C_3,3_) appear to be combined in the fast component of the bi-exponential model C_2,2_ (D_2,2_=2.2 × 10^−3^ mm^2^/s). In addition, both C_1,2_ and C_1,3_ are elevated in fatty tissues compared to the background, possibly reflecting restricted water within adipocytes. Similarly, C_2,2_ and C_2,3_ are elevated in fibroglandular tissue, but not in fat. Thus, we hypothesize that the slowest component reflects restricted diffusion signal in both tumor and fatty tissues, absent in the study by Vidić et al.

Resulting global ADCs in multi-compartment models may vary depending on the tissues included in the analysis. For example, Vidić et al showed that in a bi-exponential model the ADC of the fast component was an order of magnitude (1.89 × 10^−3^ mm^2^/s) larger than that of the slow component (3.3 × 10^−4^ mm^2^/s) when fitting cancer lesions only^23^, while the ADC values estimated here for a bi-exponential model are different by two orders of magnitude (2.2 × 10^−3^ and 2.0 × 10^−5^ mm^2^/s, respectively). We hypothesize that the difference arises from the contribution of the restricted diffusion component from fatty tissues included in control ROIs in our analysis.

In the present study, DW-MRI data acquisition between sites differ. An essential difference between sites is the imaging strategy used to decrease the magnitude of the geometric and intensity distortion artifacts that affect conventional EPI. In site 1, DW-MRI data were collected using reduced-FOV EPI in which a 2D radio-frequency pulse is used to excite spins within a reduced FOV in the phase encoding PE direction.^24^ Thus, minimizing EPI readout duration and the time during which spin dephasing occurs due to B_0_-inhomonegeities. In contrast, in site 2, parallel imaging was used to collect DW-MRI data. Parallel imaging reduces the magnitude of the distortion artifacts induced by B_0_-inhomonegeities by decreasing the number of lines collected in the PE direction. The magnitude of the distortion artifact in both strategies is reduced by the FOV reduction factor and the parallel imaging acceleration factor, both 2 in this study. The rationale to include data from different protocols, MRI scanners and vendors was to increase the generalization of the models determined here. However, the application of these models remains to be tested in an independent sample.

The T_2_-normalized (b=0 s/mm^2^) diffusion-weighted MR signal of breast lesions collected at high b-values (up to 2,500 s/mm^2^) was previously fit to bi- and tri-exponential models by Nakagawa et al. to simultaneously characterize perfusion and diffusion properties of cancer lesions.^25^ The authors reported a correlation between the tri-exponential model derived fast diffusion coefficient (D_p_, attributed to perfusion) and tumor enhancement derived from DCE-MR. In addition, the slowest (D_s_) and fastest (D_p_) diffusion coefficients (attributed to restricted diffusion and perfusion, respectively) were statistically different between malignant and benign breast lesions (D_p_ 14.81±8.30 vs. 5.54±4.64 × 10^−3^ mm^2^/s and D_s_ 0.59±0.24 vs. 0.85±0.25 ×10^−3^ mm^2^/s). Similarly, the slow diffusion coefficient estimated from a bi-exponential model was also found to be different between malignant and benign lesions (D 1.02±0.27 vs. 1.72±0.31 × 10^−3^ mm^2^/s, respectively). Direct comparison between the results of the present study and that by Nakagawa et al. is not possible due to the differences in diffusion models and lesions examined. However, results from both studies demonstrate the relevance of multi-component models in breast DW-MRI and indicate at their potential in clinical applications.

The overarching goal of this work is to generate quantitative maps in which tumor conspicuity is maximized without the use of exogenous contrast agents. The signal contribution C_2,3_ (**Figures 4 and 5**) appears to discriminate cancer tissue from control regions with minimum overlap in signal. However, visual inspection (**Figures 6 and 7**) reveals that this may not be in fact the case. Based on the plots from bi-exponential and three-component models in **Figure 5**, it becomes evident that advanced combination of all maps might be necessary to reliably maximize tumor conspicuity in breast lesions in the FOV. Future work includes the use of these RSI-derived signal contributions and advanced computer algorithms to evaluate the diagnostic value of multi-exponential models in an independent cohort. Altogether, these data may be used to aid in radiological differentiation between normal tissue, benign, and malignant breast lesions.

## Data Availability

Data may be available upon request. Please contact Dr. Dale and Dr. Rakow-Penner for further information.

## Acknowledgements

California Breast Cancer Research Program Early Career Award, GE Healthcare, NIH/NIBIB #K08EB026503, Fulbright Scholarship Program and the Research Council of Norway.

## Notes

### Competing Interest Statement

Dr. Dale reports that he was a Founder of and holds equity in CorTechs Labs, Inc., and serves on its Scientific Advisory Board. He is a member of the Scientific Advisory Board of Human Longevity, Inc. He receives funding through research grants from GE Healthcare to UCSD. Dr. Rakow-Penner is a consultant for Human Longevity, Inc. and receives funding through research grants from GE Healthcare.
The terms of these arrangements have been reviewed by and approved by UCSD in accordance with its conflict of interest policies.

